# Endovascular thrombectomy: an effective and safe therapy for perioperative ischemic stroke

**DOI:** 10.1101/2024.05.09.24307153

**Authors:** Feng Wang, Xiaoping Xu, Ling Zheng, Jiawei Zhong, En Wang, Yang Liu, Shaofa Ke

## Abstract

**Background:** Perioperative ischemic stroke is a rare but devastating complication. Mechanical thrombectomy is a promising therapeutic method, but very little data is available on its efficacy and safety. This study aims to answer this question by comparing the clinical outcomes of perioperative and community-onset stroke patients after endovascular therapy.

**Methods:** A retrospective cohort study was conducted on a total of 35 perioperative and 584 community-onset acute ischemic stroke patients who underwent endovascular thrombectomy at our hospital over the past 3.5 years. The recanalization rate, clinical recovery and cerebral hemorrhage within 90 days after therapy were compared between these two patient groups.

**Results:** Endovascular thrombectomy provided perioperative and community-onset ischemic stroke patients with comparable rates of successful reperfusion (mTICI ≥ 2b grade) (97.1% vs. 97.3%; *P* = 0. 967) and favorable functional recovery (mRS ≤ 2) (51.4% vs. 43.3%, *P* = 0.348), with no increase in severe intracranial hemorrhage (0% vs. 2.6% and 1.0%, for hematoma ≥ 30% of infarcted tissue and intraventricular hemorrhage, respectively) within 90 days. In addition, perioperative stroke patients had higher prevalence of atrial fibrillation (42.9 % vs. 26.7 %; *P* = 0.038) and intracranial cerebral artery stenosis without clear embolism (17.1 % vs. 3.8 %; *P* < 0.001) than community-onset stroke patients.

**Conclusions:** Endovascular thrombectomy is an effective and safe therapeutic approach for patients with perioperative ischemic stroke, although the results need to be validated by further studies with larger populations. Atrial fibrillation and large artery stenosis may contribute to the pathogenesis of perioperative ischemic stroke.

## Introduction

Perioperative stroke is defined as any embolic, thrombotic or hemorrhagic cerebrovascular incident within 30 days of an operation ^1^. Similar to community-onset stroke, the majority of perioperative strokes are ischemic rather than hemorrhagic ^2^. The incidence of perioperative stroke ranges from approximately 0.075% to 1.9% in non-cardiac/major vascular/neurological operations ^2–5^ to almost 10% in individuals undergoing high-risk cardiovascular surgery ^6^. In patients undergoing cardiac surgery, emboli are frequently identified as a primary cause of perioperative stroke ^1^ ^3^. In patients undergoing non-cardiac/vascular surgery, the cause of stroke is less clear. Suspected mechanisms of perioperative stroke include cerebral hypotension/low-flow states and previously undetected large-artery stenosis ^1^. It is known that intracranial atherosclerotic stenosis is more common in the Asian people than in the Western population ^7^. The mechanism of large artery stenosis in perioperative stroke should be given special attention in Chinese hospitals.

Perioperative stroke is a devastating complication for both patients and the treating surgeons/anesthesiologists ^3^ ^6^. The 30-day mortality rate for patients who suffer a perioperative stroke after non-cardiac/major-vascular/neurologic surgery is up to 8 times higher than for stroke patients unrelated to surgery ^3^. The length of hospitalization and the likelihood of discharge to a long-term care facility are also increased in patients with a perioperative stroke. An efficient therapy is urgently needed.

Intravenous thrombolysis is usually avoided in cases of perioperative stroke due to surgery itself being a contraindication for thrombolysis, leading to an elevated risk of bleeding ^2^. Mechanical endovascular thrombectomy (EVT) has become a standard treatment for stroke caused by large vessel occlusion ^8^, and also emerged as a novel therapeutic approach for perioperative stroke. Its use has been increasing over the past 15 years ^9^. A retrospective study showed that EVT can achieve comparable success in cerebral reperfusion in patients with perioperative stroke and stroke unrelated to surgery, although mortality within 3 months after EVT is higher in the former than in the latter (33.3% vs. 4.2%) ^10^. However, in a retrospective study comparing patients with in-hospital and community-onset stroke, EVT resulted in a lower recanalization rate and poorer recovery in in-hospital stroke patients ^11^. We remain optimistic about EVT therapy in perioperative stroke as surgical procedures continue to evolve and minimally invasive procedures in particular is becoming more common, which means that the pathophysiology of current perioperative stroke patients may be different from that in the past. Further research is needed to evaluate the benefits and safety of EVT in perioperative stroke.

In our research, we conducted a retrospective analysis of all 697 patients with acute ischemic stroke (AIS) who underwent EVT at our hospital over the past 3.5 years, and compared cerebral reperfusion post therapy, clinical recovery within 3 months, and incidence of cerebral bleeding between patients with perioperative and community-onset stroke. We found that EVT was an effective and safe therapeutic approach for perioperative stroke.

## Methods

### Study Design

Our project was a retrospective cohort study. The study protocol was approved by the Ethics Committee of Taizhou Hospital, Zhejiang Province, China (Registration number: K20181204). Written consent was waived due to the retrospective design. Between January 2020 and June 2023, total 697 patients aged ≥18 years suffering from AIS (including 35 patients with perioperative stroke) at our hospital received EVT according to the international and Chinese guidelines ^12^ ^13^. Perioperative stroke was defined as AIS occurring during or within 30 days following the operative procedure ^6^. Inclusion criteria were as follows: Age ≥18 years; diagnosis of acute ischemic stroke with proven arterial occlusion confirmed by computed tomography (CT) or magnetic resonance imaging (MRI); at least one attempt of EVT; EVT performed within 24 hours of symptom onset. Exclusion criteria was the presence of severe medical conditions, such as advanced heart failure, end-stage kidney disease, or terminal illness, which needed more urgent management.

### EVT and post-operative management

Endovascular therapy included stent retriever, aspiration catheter, balloon angioplasty and combination of different techniques. Balloon angioplasty was performed as the therapeutic method of first choice for recanalization in AIS patients who exhibited the “first-pass effect” of the microcatheter, in which blood was already flowing through the occluded vessel when the microcatheter was first passed and then withdrawn to the proximal side of the occluded vessel during EVT ^14^. The number of passes of catheter needed to achieve recanalization or until the end of procedure and the location of endovascular therapy were documented. At the end of EVT, the interventional neurologist determined a modified Thrombolysis in Cerebral Infarction (mTICI) score ^15^. Twenty-four hours after EVT or at any time within 24 hours when neurologic deficits progressed, patients were reexamined by head CT and scored using the National Institute of Health Stroke Scale (NIHSS). In the absence of hemorrhagic side effects, regular treatment consisted of antiplatelet agents and anticoagulants started 24 hours after EVT. Other treatment strategies included the therapy for causative surgeries in perioperative AIS patients, and statins, blood glucose and blood pressure control or combinations of these treatments according to the Chinese guidelines for the early treatment of AIS patients ^13^.

### Data Collection

The aim of this study was to compare the efficacy and safety of EVT in patients with perioperative and community-onset ischemic stroke. The primary efficacy outcome was the recovery of functional outcome as shown in the modified Rankin Scale (mRS) at 90 days after EVT. Good functional outcome was defined as mRS score ≤2. The secondary efficacy outcome was the attenuation of neurological deficits as assessed by NIHSS scores at 24 hours, and 7 days after EVT (or at the hospital discharge). The third efficacy outcome was recanalization of the occluded cerebral artery. Successful reperfusion was defined as mTICI score of 2b, 2c or 3 and unsuccessful revascularization as mTICI score of 0, 1 or 2a after EVT ^15^.

The primary safety outcome was intracranial hemorrhage as reflected by CT scanning or MRI, which were typically conducted within 7 days after EVT, with the timing determined by the individual patient’s situation and the examination capacity of our neuroradiology department. Upon neurological deterioration, a head CT was immediately performed. The Heidelberg Bleeding Classification was used to categorize the hemorrhage ^16^. The second safety outcome was all-cause mortality within 90 days after EVT.

Baseline demographic, clinical information, and laboratory findings within 24 hours after EVT were collected for all enrolled patients. Stroke subtypes were classified according to the Trial of ORG 10172 in the Acute Stroke Treatment classification (TOAST) ^17^. The Alberta stroke program early CT score (ASPECTS) was performed for both anterior and posterior circulations on the CT scan at admission ^18^ ^19^.

### Statistical analysis

The statistical analysis was conducted using SPSS software for Windows (Version 26.0, IBM, Armonk, USA). The data for continuous variables were described as mean ± SD, and categorical variables were presented as frequencies. Continuous variables were compared between independent groups by *T*-test or Mann-Whitney *U* test depending on whether the continuous variables were normally distributed. Categorical variables were compared by Pearson χ^2^ test. The association between the reduction of NIHSS score 24 hours after EVT compared with the score before EVT, and various clinical and laboratory parameters was first investigated by Pearson correlation test and then linear regression analysis using bootstrapping. *P* < 0.05 was considered statistically significant.

## Results

### Patient data

From January 2020 to June 2023, 35 perioperative AIS patients and 662 community-onset AIS patients underwent EVT in our hospital according to the international and Chinese guidelines ^12 13^. The causative surgeries of 35 perioperative stroke patients are listed in Supplementary Table 1, of whom 28 (80.0%) developed ischemic stroke minimally 1 day (1 - 28 days with a median duration of 2 days) after surgery and 10 (28.6%) had undergone cardiovascular surgery. After discharge from hospital, all AIS patients were followed up for more than 3 months by face-to-face examination or by telephone. Of community-onset AIS patients, 47 patients underwent EVT 24 hours after symptom onset due to the presence of a penumbra on CT or MRI imaging; and 31 AIS patients dropped out during the 90-day follow-up period. These 78 patients were not included in this study. Finally, 35 perioperative and 584 community-onset AIS patients were analyzed in this study (see Supplemental Figure S1, flowchart).

The demographic and baseline characteristics of perioperative and community-onset AIS patients were shown in Table 1. The atrial fibrillation was more frequent in perioperative group than in community-onset group (42.9% vs. 26.7%; χ2 (1) = 4.305, *P* = 0.038). However, both the systolic and diastolic blood pressures were lower in perioperative than in community-onset stroke patients (*P* < 0.001). The number of monocytes, concentrations of C-reactive protein and fibrinogen in the blood were higher in perioperative than in community-onset AIS patients (*P* < 0.05). Due to the causative surgeries, significantly more perioperative AIS patients than community-onset patients received anticoagulant or together with antiplatelet therapy after EVT (χ2 (3) = 54.721, *P* < 0.001). All other analyzed parameters, including age, sex, pre-stroke risk factors (e.g., hypertension, diabetes mellitus, coronary heart disease, previous stroke and current smoking), laboratory findings 24 hours after EVT (e.g., platelets, D-dimer, glucose, creatinine, cholesterol, et al.), and TOAST classification were not different between these two groups. Of note, there were no significant differences between the perioperative and community-onset groups in the neurological severity of stroke patients as shown by NIHSS scores before EVT (median value 17.4 ± 10.1 vs. 15.9 ± 7.6; Mann-Whitney-U-Test, Z = −0.553, *P* = 0.580). Thus, these 2 comparable groups of AIS patients were suitable for analysis of the efficacy and safety of EVT treatments in perioperative stroke.

**Table 1.**
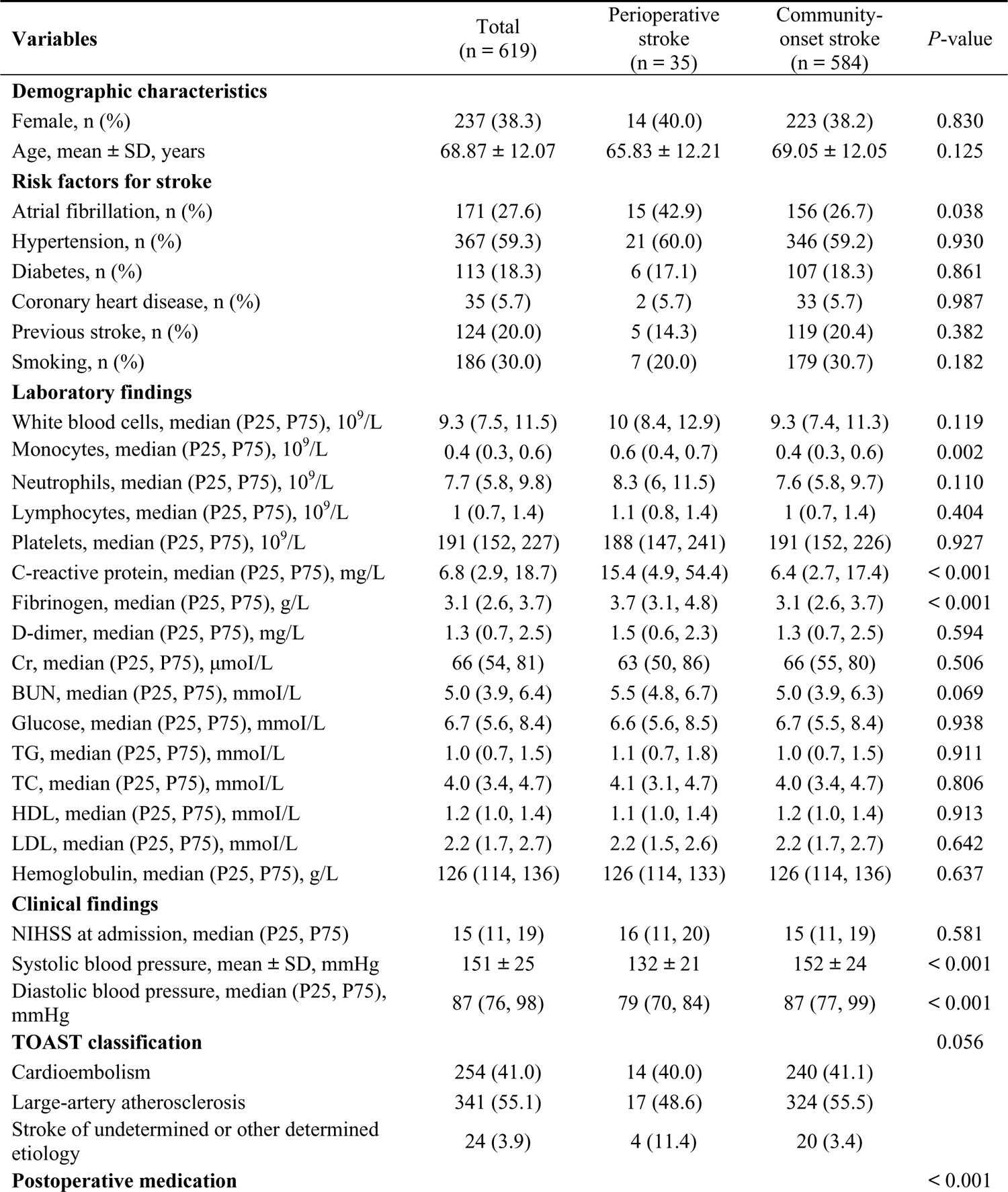
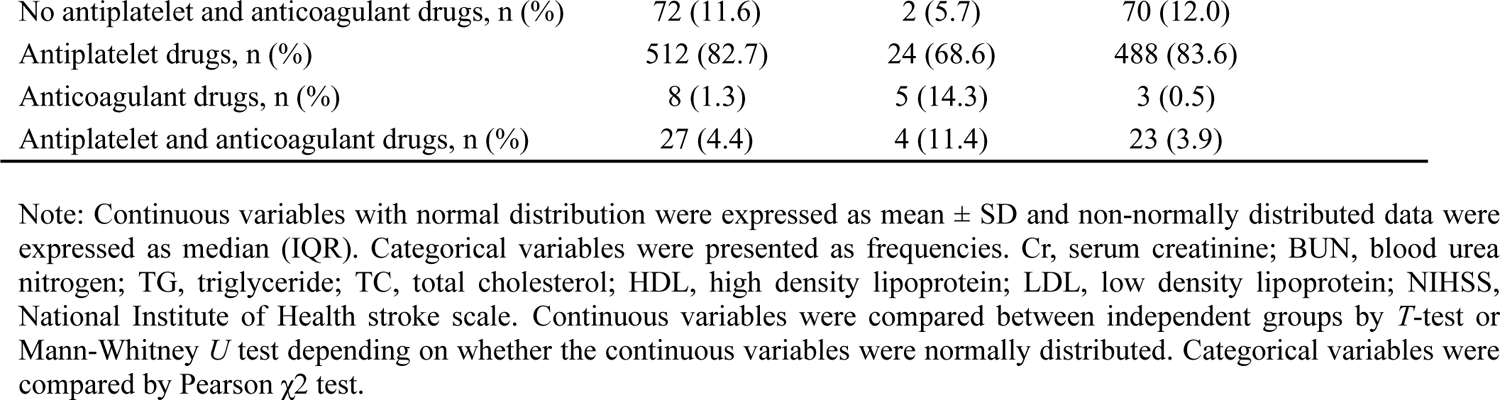
Characteristics of the patients.

| Variables | Total<br>(n = 619) | Perioperative<br>stroke<br>(n = 35) | Community-onset stroke<br>(n = 584) | P-value |
| --- | --- | --- | --- | --- |
| **Demographic characteristics** |  |  |  |  |
| Female, n (%) | 237 (38.3) | 14 (40.0) | 223 (38.2) | 0.830 |
| Age, mean ± SD, years | 68.87 ± 12.07 | 65.83 ± 12.21 | 69.05 ± 12.05 | 0.125 |
| **Risk factors for stroke** |  |  |  |  |
| Atrial fibrillation, n (%) | 171 (27.6) | 15 (42.9) | 156 (26.7) | 0.038 |
| Hypertension, n (%) | 367 (59.3) | 21 (60.0) | 346 (59.2) | 0.930 |
| Diabetes, n (%) | 113 (18.3) | 6 (17.1) | 107 (18.3) | 0.861 |
| Coronary heart disease, n (%) | 35 (5.7) | 2 (5.7) | 33 (5.7) | 0.987 |
| Previous stroke, n (%) | 124 (20.0) | 5 (14.3) | 119 (20.4) | 0.382 |
| Smoking, n (%) | 186 (30.0) | 7 (20.0) | 179 (30.7) | 0.182 |
| **Laboratory findings** |  |  |  |  |
| White blood cells, median (P25, P75), 10<sup>9</sup>/L | 9.3 (7.5, 11.5) | 10 (8.4, 12.9) | 9.3 (7.4, 11.3) | 0.119 |
| Monocytes, median (P25, P75), 10<sup>9</sup>/L | 0.4 (0.3, 0.6) | 0.6 (0.4, 0.7) | 0.4 (0.3, 0.6) | 0.002 |
| Neutrophils, median (P25, P75), 10<sup>9</sup>/L | 7.7 (5.8, 9.8) | 8.3 (6, 11.5) | 7.6 (5.8, 9.7) | 0.110 |
| Lymphocytes, median (P25, P75), 10<sup>9</sup>/L | 1 (0.7, 1.4) | 1.1 (0.8, 1.4) | 1 (0.7, 1.4) | 0.404 |
| Platelets, median (P25, P75), 10<sup>9</sup>/L | 191 (152, 227) | 188 (147, 241) | 191 (152, 226) | 0.927 |
| C-reactive protein, median (P25, P75), mg/L | 6.8 (2.9, 18.7) | 15.4 (4.9, 54.4) | 6.4 (2.7, 17.4) | < 0.001 |
| Fibrinogen, median (P25, P75), g/L | 3.1 (2.6, 3.7) | 3.7 (3.1, 4.8) | 3.1 (2.6, 3.7) | < 0.001 |
| D-dimer, median (P25, P75), mg/L | 1.3 (0.7, 2.5) | 1.5 (0.6, 2.3) | 1.3 (0.7, 2.5) | 0.594 |
| Cr, median (P25, P75), μmol/L | 66 (54, 81) | 63 (50, 86) | 66 (55, 80) | 0.506 |
| BUN, median (P25, P75), mmol/L | 5.0 (3.9, 6.4) | 5.5 (4.8, 6.7) | 5.0 (3.9, 6.3) | 0.069 |
| Glucose, median (P25, P75), mmol/L | 6.7 (5.6, 8.4) | 6.6 (5.6, 8.5) | 6.7 (5.5, 8.4) | 0.938 |
| TG, median (P25, P75), mmol/L | 1.0 (0.7, 1.5) | 1.1 (0.7, 1.8) | 1.0 (0.7, 1.5) | 0.911 |
| TC, median (P25, P75), mmol/L | 4.0 (3.4, 4.7) | 4.1 (3.1, 4.7) | 4.0 (3.4, 4.7) | 0.806 |
| HDL, median (P25, P75), mmol/L | 1.2 (1.0, 1.4) | 1.1 (1.0, 1.4) | 1.2 (1.0, 1.4) | 0.913 |
| LDL, median (P25, P75), mmol/L | 2.2 (1.7, 2.7) | 2.2 (1.5, 2.6) | 2.2 (1.7, 2.7) | 0.642 |
| Hemoglobin, median (P25, P75), g/L | 126 (114, 136) | 126 (114, 133) | 126 (114, 136) | 0.637 |
| **Clinical findings** |  |  |  |  |
| NIHSS at admission, median (P25, P75) | 15 (11, 19) | 16 (11, 20) | 15 (11, 19) | 0.581 |
| Systolic blood pressure, mean ± SD, mmHg | 151 ± 25 | 132 ± 21 | 152 ± 24 | < 0.001 |
| Diastolic blood pressure, median (P25, P75), mmHg | 87 (76, 98) | 79 (70, 84) | 87 (77, 99) | < 0.001 |
| **TOAST classification** |  |  |  | 0.056 |
| Cardioembolism | 254 (41.0) | 14 (40.0) | 240 (41.1) |  |
| Large-artery atherosclerosis | 341 (55.1) | 17 (48.6) | 324 (55.5) |  |
| Stroke of undetermined or other determined etiology | 24 (3.9) | 4 (11.4) | 20 (3.4) |  |
| **Postoperative medication** |  |  |  | < 0.001 |

|  |  |  |  |
| --- | --- | --- | --- |
| No antiplatelet and anticoagulant drugs, n (%) | 72 (11.6) | 2 (5.7) | 70 (12.0) |
| Antiplatelet drugs, n (%) | 512 (82.7) | 24 (68.6) | 488 (83.6) |
| Anticoagulant drugs, n (%) | 8 (1.3) | 5 (14.3) | 3 (0.5) |
| Antiplatelet and anticoagulant drugs, n (%) | 27 (4.4) | 4 (11.4) | 23 (3.9) |

Note: Continuous variables with normal distribution were expressed as mean ± SD and non-normally distributed data were expressed as median (IQR). Categorical variables were presented as frequencies. Cr, serum creatinine; BUN, blood urea nitrogen; TG, triglyceride; TC, total cholesterol; HDL, high density lipoprotein; LDL, low density lipoprotein; NIHSS, National Institute of Health stroke scale. Continuous variables were compared between independent groups by *T*-test or Mann-Whitney *U* test depending on whether the continuous variables were normally distributed. Categorical variables were compared by Pearson $\chi^2$ test.

### EVT provided perioperative AIS patients with a comparable reperfusion rate and 3-month rehabilitation, and even a faster recovery within 24 hours compared to community-onset AIS patients

There was no significant difference in the rate of successful reperfusion [mTICI ≥ 2b grade]) between perioperative and community-onset AIS patients (Table 2; 97.1% vs. 97.3%; χ2 (1) = 0.002, *P* = 0.967). Similarly, the distribution of mTICI scores (1 = mTICI 0, 1 or 2a, 2 = mTICI 2b, and 3 = mTICI 3), which categorize the level of successful reperfusion, did not show any significant differences between these two AIS patient groups (Supplementary Fig. S2; χ2 (2) = 0.035, *P* = 0.983). It was also found that the causative surgeries did not complicate the EVT procedure, as the number of catheter passes required for recanalization did not vary significantly between these two groups of patients (Table 2; 1.6 ± 0.9 vs. 1.4 ± 0.8; Mann-Whitney-U-Test, Z = −1.436, *P* = 0.151). In addition, perioperative and community-onset AIS patients did not differ in operative time of EVT and the site of arterial occlusion (Table 2).

**Table 2.** Characteristics of endovascular therapy procedure.

| Variables | Total<br>(n = 619) | Perioperative<br>stroke<br>(n = 35) | Community-onset stroke<br>(n = 584) | P-value |
| --- | --- | --- | --- | --- |
| **ASPECT score**, median (P25, P75) | 9 (8, 10) | 10 (10, 10) | 9 (8, 10) | < 0.001 |
| **Fibrinolysis** | 189 (30.5) | 2 (5.7) | 187 (32.0) | 0.001 |
| **Endovascular therapy** |  |  |  |  |
| Onset-to-puncture time, median (P25, P75), minutes | 420 (300, 655) | 239 (178, 299) | 433 (315, 666) | < 0.001 |
| Puncture-to-recanalization time, median (P25, P75), minutes | 66 (45, 100) | 60 (38, 90) | 66 (45, 102) | 0.129 |
| Successful reperfusion (mTICI ≥ 2b grade) | 602 (97.3) | 34 (97.1) | 568 (97.3) | 0.967 |
| Numbers of passes of catheter, median (P25, P75) | 1 (1, 2) | 1 (1, 2) | 1 (1, 2) | 0.151 |
| **Location of occluded artery** |  |  |  | 0.222 |
| Terminus internal carotid artery, n (%) | 114 (18.4) | 10 (28.6) | 104 (17.8) |  |
| Middle cerebral artery, n (%) | 316 (51.1) | 20 (57.1) | 296 (50.7) |  |
| Anterior cerebral artery, n (%) | 16 (2.6) | 1 (2.9) | 15 (2.6) |  |
| ≥ 2 arteries in anterior circulation, n (%) | 58 (9.4) | 0 (0) | 58 (9.9) | |
| Basilar artery, n (%) | 80 (12.9) | 3 (8.6) | 77 (13.2) |  |
| Vertebral artery, n (%) | 25 (4.0) | 0 (0) | 25 (4.3) |  |
| Posterior cerebral artery, n (%) | 10 (1.6) | 1 (2.9) | 9 (1.5) |  |
| **Endovascular therapy procedure** |  |  |  | < 0.001 |
| Stent retriever, n (%) | 423 (68.3) | 24 (68.6) | 399 (68.3) |  |
| Balloon angioplasty, n (%) | 28 (4.5) | 6 (17.1) | 22 (3.8) |  |
| Stent retriever and balloon angioplasty, n (%) | 127 (20.5) | 1 (2.9) | 126 (21.6) |  |
| Aspiration catheter, n (%) | 17 (2.7) | 3 (8.6) | 14 (2.4) |  |
| Aspiration catheter or stent retriever, n (%) | 24 (3.9) | 1 (2.9) | 23 (3.9) |  |
| **Clinical findings** |  |  |  |  |
| NIHSS at 24 hours after EVT, median (P25, P75) | 13 (7, 19) | 8 (5, 18) | 13 (7, 19) | 0.094 |
| Reduction of NIHSS at 24 hours compared to NIHSS before EVT, median (P25, P75) | 0 (0, 5) | 5 (0, 10) | 0 (0, 4) | < 0.001 |
| NIHSS at 7 days after EVT, median (P25, P75) | 7 (2, 15) | 3 (1, 14) | 7 (2, 15) | 0.204 |
| mRS ≤ 2 at 90 days after EVT, n (%) | 271 (43.8) | 18 (51.4) | 253 (43.3) | 0.348 |
| Mortality at the end of study, n (%) | 108 (17.4) | 4 (11.4) | 104 (17.8) | 0.933 |

Note: Non-normally distributed data were expressed as median (IQR). Categorical variables were presented as frequencies. Continuous variables were compared between independent groups by Mann-Whitney *U* test. Categorical variables were compared by Pearson $\chi^2$ test.

It should be noted that there were more patients in the perioperative stroke group than in the community-onset stroke group (Table 2; 17.1% vs. 3.8%; χ2 (1) = 13.680, *P* < 0.001), who had the “first-pass effect” of the microcatheter, which suggests severe stenosis in the intracranial cerebral arteries and not severe embolism-related vascular occlusion ^20^, and whose cerebral blood flow could be sufficiently improved by balloon angioplasty alone.

Possibly due to the professional medical care in the hospital, patients with perioperative stroke were much easier to recognize than patients with community-onset stroke. As shown in Table 2, the time from onset of symptoms to puncture was significantly shorter (202.5 ± 217.3 vs. 448.6 ± 279.7 minutes; Mann-Whitney-U-Test, Z = −6.553, *P* < 0.001), and the ASPECT score was significantly higher (9.9 ± 0.7 vs. 8.5 ± 1.8; Mann-Whitney-U-Test, Z = −5.454, *P* < 0.001) in perioperative AIS patients than in the community-onset AIS controls. The improvement in neurological deficits shown by the reduction in NIHSS score 24 hours after EVT compared with NIHSS score before EVT was significantly more pronounced in perioperative stroke patients than in community-onset stroke patients (Table 2; ΔNIHSS score: 5.5 ± 8.9 vs. 1.56 ± 6.0; Mann-Whitney-U-Test, Z = −3.052, *P* = 0.002). Unfortunately, this favorable recovery after EVT was lost in perioperative AIS patients compared to community-onset AIS patients in the following 7 days, as the NIHSS scores did not differ between perioperative and community-onset AIS patients 7 days after EVT (Table 2; 9.4 ± 9.4 vs. 11.0 ± 12.0; Mann-Whitney-U-Test, Z = −1.271, *P* = 0.204).

To investigate the underlying mechanisms hindering further recovery of perioperative AIS patients, we combined perioperative and community-onset AIS patients and performed a correlation analysis to identify variables related to NIHSS score reduction within 24 hours after EVT (see Supplementary Table 2). We then performed a regression analysis with NIHSS reduction as the dependent variable and the correlated parameters as independent variables. We found that perioperative stroke classification, smoking and NIHSS score at admission positively predicted, while diabetes, white blood cell count, D-dimer, creatine, systolic blood pressure, puncture-to-recanalization time and location of occluded artery with MCA as the reference location negatively predicted NIHSS score reduction (Supplementary Table 3). In combination with the high levels of monocytes and C-reactive protein in perioperative AIS patients compared to community-onset stroke patients (Table 1), inflammatory activation could be an influential factor affecting clinical recovery.

To assess the functional recovery of all AIS patients under study, mRS scores were evaluated 90 days after EVT. The analysis revealed no significant differences in the distribution of mRS scores between patients with perioperative and community-onset AIS (Figure 1; χ2 (6) = 5.923, *P* = 0.432). The percentage of patients achieving a favorable outcome (mRS ≤ 2) in perioperative group was comparable with that in community-onset AIS group (Table 2; 51.4% vs. 43.3%, χ2 (1) = 0.882, *P* = 0.348). The mortalities during the entire course of the study were also not significantly different between perioperative and community-onset AIS patients (Table 2; 11.4% vs. 17.8%, χ2 (1) = 0.933, *P* = 0.334).

**Figure 1.**
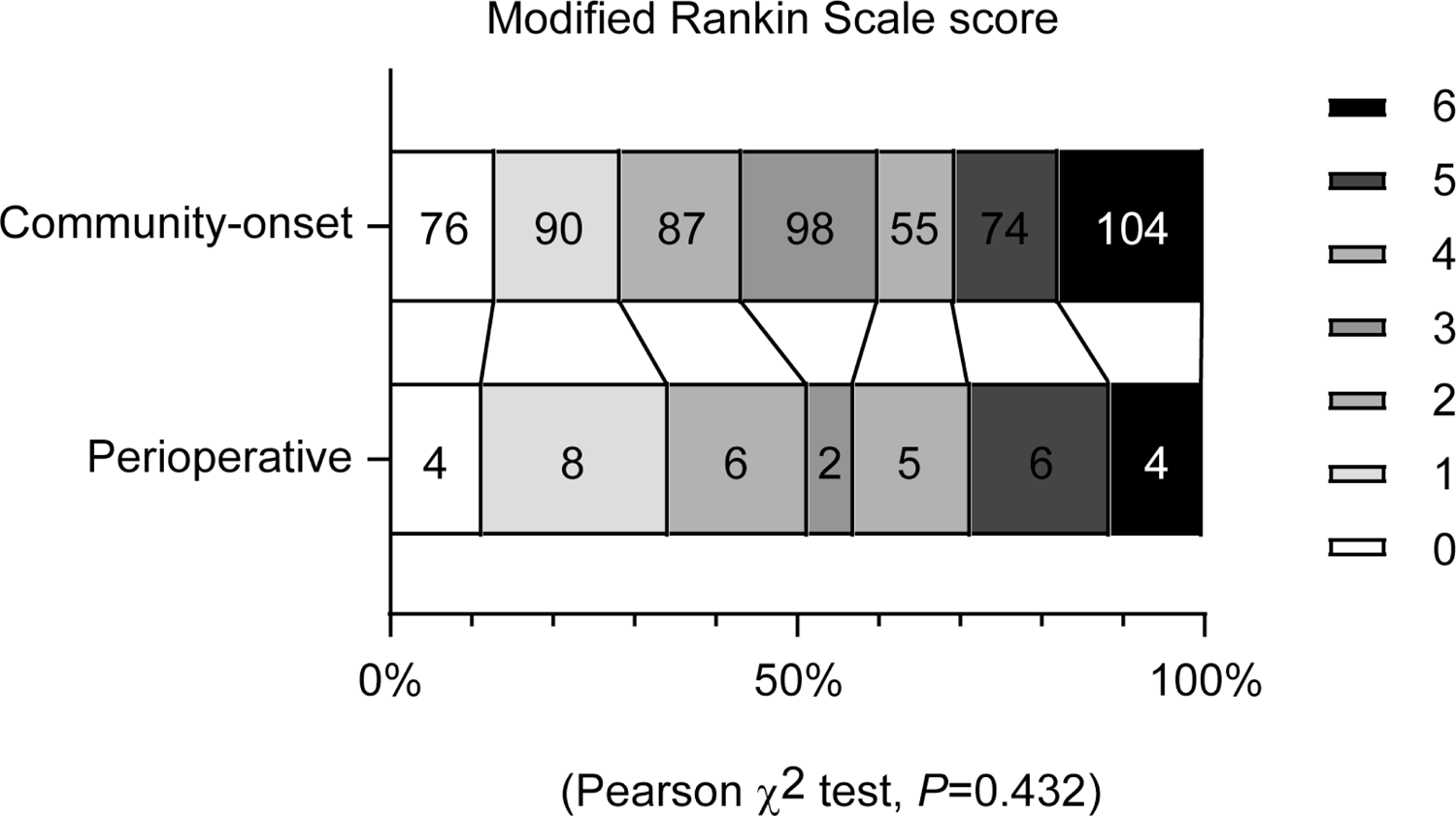
Endovascular thrombectomy achieves comparable functional recovery in perioperative and community-onset stroke patients. Functional recovery of patients with acute ischemic stroke was assessed using the modified Rankin Scale (mRS) at 90 days after endovascular thrombectomy. The mRS scores range from 0 to 6, with score 0 indicating no symptoms, to score 5 indicating severe disability and score 6 indicating death. The distribution of patients with different mRS scores did not differ between the perioperative and community-onset stroke patient groups. χ^2^ test, n = 35 and 584 for perioperative and community-onset stroke groups, respectively.

### EVT did not increase a hemorrhagic risk for perioperative AIS patients compared with community-onset AIS patients

Hemorrhagic transformation is the most important adverse event after EVT therapy in AIS patients. The percentage of patients diagnosed with overall intracranial hemorrhage within 7 days after EVT was comparable in the perioperative and community-onset AIS groups (Table 3; 25.7% vs. 19.5%; χ2 (1) = 0.796, *P* = 0.372). After dividing the AIS patients into subgroups according to the Heidelberg Bleeding Classification ^16^, we found that perioperative and community-onset AIS patients differed significantly in terms of intracranial hemorrhage (Table 3; χ2 (5) = 16.299, *P* = 0.004). However, the difference was limited in the subgroups of HI1 (11.4% vs. 3.8; χ2 (1) = 4.817, *P* = 0.028) and PH1-type (8.6% vs. 2.4%; χ2 (1) = 4.713, *P* = 0.030) hemorrhages, and subarachnoid hemorrhage (5.7% vs. 0.9%; χ2 (1) = 6.970, *P* = 0.008) (Table 3). There were no perioperative AIS patients in the subgroups of PH2 and intraventricular hemorrhage, perhaps due to the limited sample size (Table 3).

**Table 3.** Characteristics of intracranial hemorrhage.

| Variables | Total<br>(n = 619) | Peri-operative<br>stroke<br>(n = 35) | Community-onset stroke<br>(n = 584) | $\chi^2$ test<br>*P*-value |
| --- | --- | --- | --- | --- |
| **Intracranial hemorrhages** | 123 (19.9) | 9 (25.7) | 114 (19.5) | 0.372 |
| **The Heidelberg Bleeding Classification** |  |  |  | 0.004 |
| 0. No hemorrhages | 496 (80.1) | 26 (74.3) | 470 (80.5) |  |
| 1. Hemorrhagic transformation of infarcted brain tissue |  |  |  |  |
| 1a (HI1), Scattered small petechiae, no mass effect | 26 (4.2) | 4 (11.4) | 22 (3.8) |  |
| 1b (HI2), Confluent petechiae, no mass effect | 54 (8.7) | 0 (0) | 52 (8.9) |  |
| 1c (PH1), Hematoma within infarcted tissue, occupying <30%, no substantive mass effect | 16 (2.6) | 3 (8.6) | 14 (2.4) |  |
| 2. Intracerebral hemorrhage within and beyond infarcted brain tissue |  |  |  |  |
| PH2, Hematoma occupying 30% or more of the infarcted tissue, with obvious mass effect | 14 (2.3) | 0 (0) | 15 (2.6) |  |
| 3. Intracerebral hemorrhage outside the infarcted brain tissue |  |  |  |  |
| 3a Parenchymal hematoma remote from infarcted brain tissue | 13 (2.1) | 0 (0) | 1 (0.2) |  |
| 3b Intraventricular hemorrhage |  | 0 (0) | 6 (1.0) |  |
| 3c Subarachnoid hemorrhage |  | 2 (5.7) | 5 (0.9) |  |

## Discussion

Intravenous thrombolysis is often not feasible in perioperative AIS patients as it increases the risk of bleeding ^2^ ^6^. EVT is the most useful method for recanalization of cerebral arteries in perioperative AIS patients and is increasingly practiced in many hospitals ^9^. However, the efficacy of EVT in improving patient outcomes remains uncertain. Our study showed that EVT in perioperative AIS patients resulted in a similar reperfusion rate and 3-month recovery without an increased risk of bleeding and death compared to patients with community-onset stroke.

In our study, we used the entire cohort of community-onset stroke patients instead of selecting certain matched AIS patients as a control group for perioperative AIS patients, which we believe can limit the sampling bias and better represent the real-world information. The NIHSS score is higher in perioperative AIS patients than in community-onset AIS patients, even though there is no statistical difference, which demonstrates the efficacy of EVT in perioperative stroke. We found that atrial fibrillation was significantly more common in perioperative AIS patients than in community-onset AIS patients, which is consistent with previous studies that perioperative or postoperative atrial fibrillation was associated with an increased risk of both early and long-term ischemic stroke, especially in patients undergoing non-cardiac surgery ^21^ ^22^. Our study seems to show that undetected stenosis of the intracranial arteries is a pathogenic mechanism of perioperative stroke. We observed more AIS patients in the perioperative stroke group than in the community-onset stroke group (17.1 % vs. 3.8 %) who had atherosclerotic stenosis in the cerebral arteries without clear thrombi or emboli, in whom balloon angioplasty alone was able to restore blood flow to the brain tissue. Intracranial atherosclerotic stenoses pose a challenge for EVT in AIS patients as they increase intraprocedural re-occlusion ^23^. A recent study in AIS patients with intracranial atherosclerosis-related large vessel occlusion showed that balloon angioplasty as a first-choice recanalization strategy has a higher efficiency in recanalization and better functional outcomes at 90 days compared to thrombectomy ^14^.

Our study showed that the recanalization rate after EVT did not differ between perioperative and community-onset AIS stroke patients, which corroborates previous studies on stroke patients after both cardiovascular and non-cardiovascular surgeries ^10^ ^24^. Importantly, the time from symptom onset to groin puncture in our study is shorter in perioperative stroke patients than in community-onset AIS stroke patients (median time, 239 vs. 433 minutes). The symptom onset-to-groin puncture time is strongly associated with better clinical outcome, e.g., functional independence at discharge ^25^. We found that the reduction in NIHSS scores within 24 hours of EVT was significantly greater in perioperative AIS patients than in community-onset AIS patients, although this difference disappeared when patients were discharged from our hospital. We observed that: 1) the increase in white blood cell count was negatively associated with NIHSS score reduction after EVT; and 2) monocyte counts and C-reactive protein amount were higher in perioperative patients than in community-onset AIS patients. It therefore stands to reason that inflammatory activation impedes functional recovery in perioperative AIS patients. It has been reported that the increase in C-reactive protein ^26^, or the inflammatory index C-reaction protein × neutrophil/lymphocyte count ^27^ predicts an unfavorable functional outcome in AIS patients 90 day after EVT.

Our study also showed that EVT provided perioperative and community-onset AIS patients with comparable functional recovery and mortality 3 months after EVT, which is different from a previous observation that perioperative AIS patients had a higher rate of death within 3 months of EVT than community-onset AIS patients ^10^. Not surprisingly, the underlying diseases requiring surgeries and comorbidities influence the outcome of perioperative AIS patients after EVT. In the previous study ^10^, there were 68% perioperative AIS patients receiving cardiovascular surgery and 12% receiving neurosurgery, while we had only 29% patients in the perioperative AIS group, who had undergone cardiovascular procedure. In studies of the therapeutic efficacy of EVT in in-hospital and community-onset AIS patients, the former generally had poorer recovery and higher mortality, which was correlated with the modified Charlson Comorbidity Index (mCCI), incorporating 7 comorbidities, age, diabetes, anemia, active cancer, myocardial infarction, congestive heart disease, and ulcer disease into the model ^11^ ^28^. We further suppose that the widely used mini-invasive surgery in our studying cohort may favor the functional recovery of perioperative AIS patients. Compared to open surgical procedures such as sternotomy or open thoracotomy, minimally invasive techniques offer advantages to patients, such as a lower risk of surgical and postoperative complications, shorter recovery times and a reduction in postoperative pain, which can reduce systemic inflammation and hemodynamic changes ^29^.

Consistent with previous studies ^10^ ^11^ ^24^ ^28^, EVT did not lead to an increase in total intracranial hemorrhage in perioperative AIS patients compared to community-onset AIS patients. However, when hemorrhages with different subtypes according to the Heidelberg hemorrhage classification were considered, there were significantly more perioperative AIS patients with HI1 and PH1 type hemorrhages and subarachnoid hemorrhage than community-onset AIS patients. Nevertheless, these types of intracranial hemorrhage were generally thought to have little impact on patient outcome. There were no perioperative AIS patients with PH2-type hemorrhage and intravascular hemorrhage, which often lead to symptomatic intracranial hemorrhage and poorer prognosis ^16^ ^30^.

Obviously, our study has a limitation. The study population was recruited from a single institution with a limited number of patients. The patients cannot be further subdivided to evaluate the therapeutic efficacy and safety of EVT in perioperative AIS patients with cardiovascular or non-cardiovascular procedures. The incidence of PH2-type intracranial hemorrhage and intravascular hemorrhage in perioperative AIS patients after EVT also remains unclear.

## Conclusion

Our study shows that endovascular thrombectomy or balloon angioplasty is an effective and safe therapeutic method for the treatment of perioperative strokes with large vessel occlusions. It is helpful for clinicians to make treatment decisions for perioperative stroke patients. Our study also supports the hypothesis that atrial fibrillation and intracranial cerebral artery stenosis contribute to the occurrence of perioperative stroke. However, our results need to be validated by further studies with larger populations.

## Data Availability

The data that support the findings of this study are available on request from the corresponding author, Y.L. and S.K. The data are not publicly available because it contains information that could compromise the privacy of the research participants.

## Acknowledgments

The authors thank all patients for contributing to this study.

## Funding

This work was supported by grants from Zhejiang Provincial Basic and Public Welfare Research Program (No. GF21H020023); Zhejiang Provincial Medicine and Health Research Foundation (grant number: 2021RC141).

## Contributions

Study idea and design: Feng Wang, Yang Liu and Shaofa Ke; Coordination and organization: Feng Wang and Shaofa Ke; Participants enrollment: En Wang; Data collection and statistical analysis: Xiaoping Xu, Ling Zheng, and Yang Liu; Writing, editing and revision: Feng Wang and Yang Liu.

## Ethics declarations Conflict of interest

The author(s) declared no potential conflicts of interest with respect to the research, authorship, and/or publication of this article.

## Ethical standards

The studies involving human participants were reviewed and approved by the Ethics Committee of Taizhou Hospital, Zhejiang Province, China (K20181204).

## Notes

### Competing Interest Statement

The authors have declared no competing interest.

